# Automated Deep Learning Pipeline for Characterizing Left Ventricular Diastolic Function

**DOI:** 10.1101/2025.04.29.25326683

**Authors:** Victoria Yuan, Yuki Sahashi, Hirotaka Ieki, Miloš Vukadinovic, Christina Binder, Konrad Pieszko, Andrew P. Ambrosy, Paul P. Cheng, Susan Cheng, David Ouyang

## Abstract

**Introduction:** Left ventricular diastolic dysfunction (LVDD) is most commonly evaluated by echocardiography. However, without a sole identifying metric, LVDD is assessed by a diagnostic algorithm relying on secondary characteristics that is laborious and has potential for interobserver variability.

**Methods:** To characterize concordance in clinical evaluations of LVDD, we evaluated historical echocardiogram studies at two academic medical centers for variability between clinician text reports and assessment by 2016 American Society of Echocardiography (ASE) guidelines. We then developed a workflow of 8 artificial intelligence (AI) models trained on over 155,000 studies to automate assessment of LVDD. Model performance was evaluated on temporally distinct held-out test sets from two academic medical centers.

**Results:** In a validation cohort of 955 studies from Cedars-Sinai Medical Center, our AI workflow demonstrated 76.5% agreement and a weighted Cohen’s kappa of 0.52 with ASE guideline assessment using human measurements. In contrast, the clinician report evaluation had 48.5% agreement and a weighted Cohen’s kappa of 0.29 with ASE guidelines. In the Stanford Healthcare cohort of 1,572 studies, the AI workflow had 66.7% agreement and a weighted Cohen’s kappa of 0.27 with ASE guidelines, while the clinician assessment had 32.7% agreement and a weighted Cohen’s kappa of 0.06. Performance was consistent across patient subgroups stratified by sex, age, hypertension, diabetes, obesity, and coronary artery disease.

**Conclusion:** Clinicians are often inconsistent in evaluating LVDD. We developed an AI pipeline that automates the clinical workflow of grading LVDD, which can contribute to improved diagnosis of heart failure.

## Introduction

Left ventricular diastolic dysfunction (LVDD) results from impaired LV relaxation and increased chamber stiffness, which can lead to increased filling pressures. LVDD develops early in cardiac pathologies. Thus, characterizing LVDD is integral to diagnosis and prognosis for a range of cardiac and extra-cardiac diseases^1–6^ including evaluating dyspnea and risk stratifying heart failure, end-stage renal disease, coronary artery disease, myocardial infarction, and other pathologies^7,8^. However, evaluation of LVDD is not always reliable or consistent.

LVDD is evaluated by echocardiography using guidelines from the American Society of Echocardiography (ASE) outlined in 2016^10^. However, this diagnostic algorithm is time-consuming and extensive, requiring integration of multiple parameters, including annular velocities, mitral valve inflow, tricuspid regurgitation, and left atrial volume. In general clinical practice, LVDD is often missed, with up to 75% of patients missing measurements for diastology^10–13^, which can lead to missed diagnoses of heart failure with preserved ejection fraction (HFpEF) and imprecise generalized assessment of diastolic function^11^. Moreover, the individual parameters required for assessing diastolic function have high interobserver variability^14,15^, leading to variability in the assessment of diastolic function^16^.

Deep learning (DL) applied to echocardiography has shown promise in facilitating reproducible, efficient assessments and measurements^16–21^. DL models for both electrocardiograms as well as single view echocardiogram videos have been developed to assess LVDD holistically, leading to black-box assessments not directly related to measurements^23,24^. Models have been developed for individual measurements in the workflow of assessing LVDD^19,24,25^; however its impact on clinical practice and overall diagnosis has not been well assessed. Creating a DL pipeline with individual models trained on large datasets for each individual task in the workflow of assessing LVDD can offer a robust method to automate guideline-based diastology assessment.

In this work, we develop a DL pipeline consisting of 9 models to automate the clinical workflow for grading LVDD. Our DL workflow includes view classification, measurement of B-mode and Doppler parameters, and evaluation of diastolic function according to the 2016 ASE guidelines. Our pipeline demonstrated strong concordance with assessment of LVDD using manual clinical measurements in two distinct patient cohorts, and show this performance improves significantly upon historical clinician reports.

## Methods

### Investigating Clinician Concordance in Evaluating Diastology

We compared diastolic function grade defined by 2016 ASE guidelines with the clinical echocardiography report assessment of diastolic function in 124,524 studies across 87,425 unique CSMC patients and 1,572 studies across 1,560 SHC patients after April 2016. Limited studies with less than 2 parameters for diastology were excluded. Patients were excluded based on history of mitral valve pathology, mitral valve repair, tachycardia during diastolic evaluation, atrial fibrillation and other arrhythmias, pacemaker implantation, history of orthotopic heart transplant, or if it was a limited echocardiographic study. We extracted clinician evaluations of diastology from final text reports, with clinician assessments of mild LVDD mapped to grade 1 diastolic dysfunction, moderate to grade 2, and severe to grade 3.

### Deep Learning Workflow to Automate Diastology

Our DL pipeline automates the clinical workflow for diastolic function by: 1) Classifying the view of each echocardiography image 2) Identifying and removing low-quality files unsuitable for downstream analysis 3) Measuring the necessary parameters for diastology with DL models 4) Grades diastolic function using the 2016 ASE guidelines (Figure 1). Multi-view classification was conducted with a previously validated model from EchoPrime and quality control with a model from EchoNet-Measurements^19,26^.

**Figure 1.**
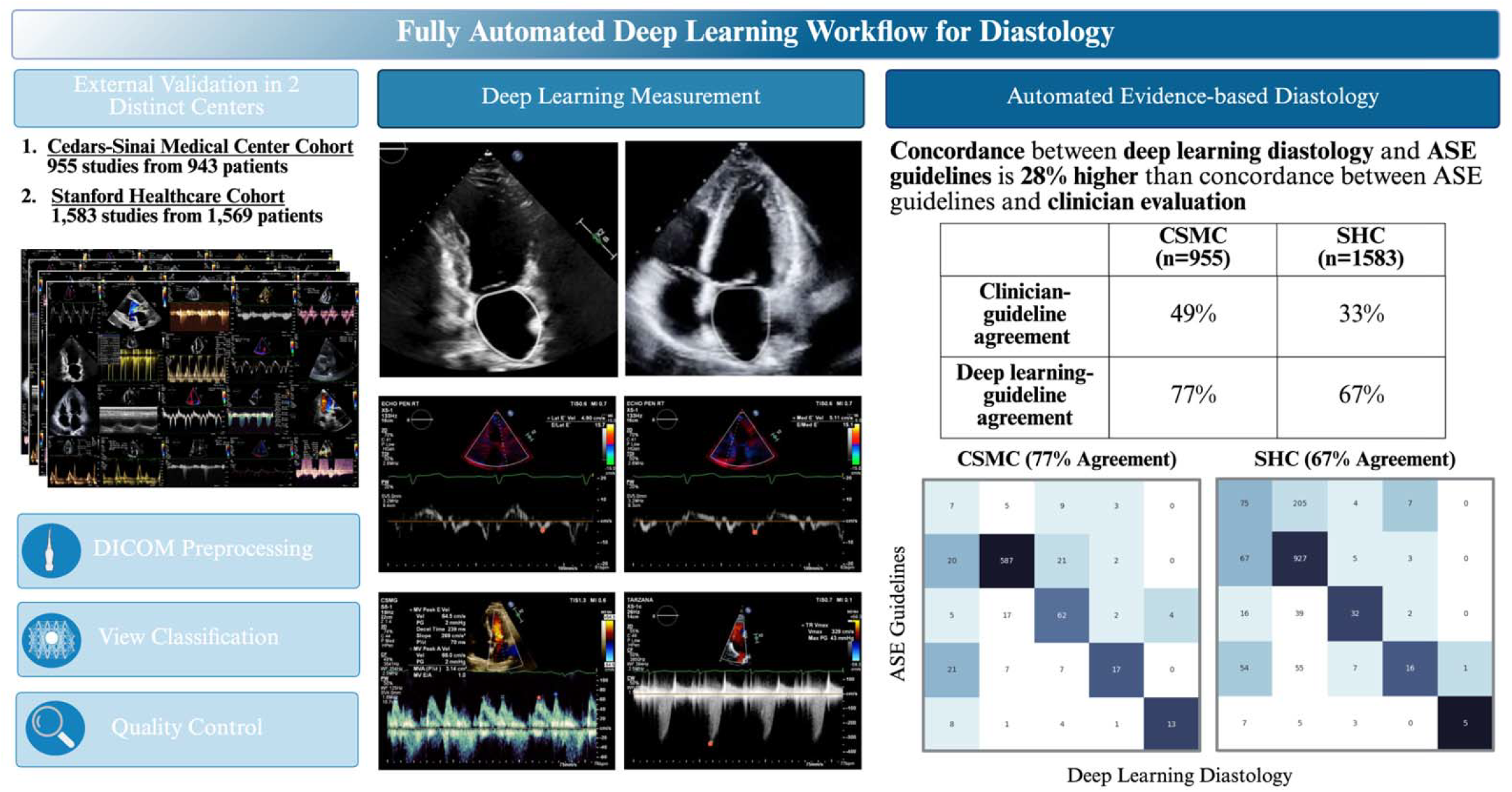
Schematic of deep learning-based pipeline that automates the clinical workflow for diastolic function

The parameters for diastology were derived using individual DL models. First, left ventricular ejection fraction (LVEF) was calculated using EchoNet-Dynamic^17^. Doppler parameters for medial and lateral e’ velocities, mitral valve (MV) peak E and A velocities, and maximum tricuspid regurgitant velocity (TR V_max_) were calculated with EchoNet-Measurements^20^. MV E/e’ was derived as MV E divided by the average of medial e’ and lateral e’ velocities. Finally, we employed DeepLabv3 to segment the left atrium (LA) from apical 4-chamber (A4C) and apical 2-chamber (A2C) echocardiogram videos. The frame with the maximum LA area corresponded to LA diastole and utilized to calculate left atrial volume (LAV). When both A4C and A2C views were available, LAV was calculated with the biplane method of discs, otherwise via single plane method of discs. Patient body surface area (BSA) was calculated from height and weight.

### External Validation of DL Workflow for Diastolic Function

We deployed our workflow to automatically characterize diastology in two patient cohorts – a temporally distinct cohort of CSMC patients who received care from June 2022 to November 2024 and a geographically distinct cohort from SHC from April 2016 to August 2018. Patients with history of mitral valvular pathology, orthotopic heart transplant, atrial fibrillation and other arrhythmias, and tachycardia at the time of assessment for diastolic dysfunction were excluded. We then compared the results of our DL diastology against diastology extracted from clinician reports and diastology calculated from clinical echocardiographic measurements based on ASE guidelines. Patients without LAV measured on echocardiography but with clinician text reports noting LA enlargement were classified as having abnormal LAV_i_ ≥ 34 mL/m^2^.

### Statistical Analysis

Concordance was quantified with percent agreement, quadratic-weighted Cohen’s kappa, and confusion matrices. Finally, we divided patients into subgroups based on sex, age ≥65, hypertension, diabetes mellitus, history of coronary artery disease, and obesity with body mass index (BMI) ≥ 30. Additionally, we analyzed the ability of DL diastology to identify elevated LV filling pressures according to ASE guidelines; patients with grade 2 or 3 diastolic dysfunction had elevated filling pressure whereas patients with normal or grade 1 LVDD did not.

### Data and Code Availability

The code and model weights will be available on GitHub at https://github.com/echonet/diastology. Patient data will not be available due to its potentially identifiable nature.

## Results

### Evaluating Clinician Variability in Diastology Assessment

We compared evaluations of diastology in 124,524 studies across 87,425 CSMC patients and 1,572 studies across 1,560 SHC patients (Table 1, Supplemental Table 1). Diastolic function was compared between two sources – extracted from final echocardiography text report and calculated from clinical echocardiography parameters according to ASE guidelines. Clinician assessments of diastolic function had only 30.8% agreement and 32.7% agreement with ASE guidelines in the CSMC and SHC cohorts, respectively (Figure 2, Figure 3b). The weighted Cohen’s kappa *κ* between clinician reports and ASE guidelines was 0.20 at CSMC and 0.06 at SHC.

**Table 1.** Demographics and comorbidities of CSMC and SHC validation cohorts.

| **Characteristic** | **CSMC (n=955)** | **SHC (n=1572)** |
| --- | --- | --- |
| Female | 448 ( 46.9%) | 809 (51.1%) |
| Male | 507 (53.1%) | 773 (48.8%) |
| Age (mean±std) | 63.9±17.2 | 58.6±18.1 |
| Race and Ethnicity |  |  |
| American Indian / Alaskan Native | 2 (0.2%) | 4 (0.3%) |
| Asian | 90 (9.4%) | 338 (21.5%) |
| Black / African-American | 133 (13.9%) | 89 (5.7%) |
| Hispanic | 116 (12.4%) | 184 (11.7%) |
| Native Hawaiian / Pacific Islander | 6 (0.6%) | 25 (1.6%) |
| White | 601 (62.9%) | 742 (47.2%) |
| Unknown | 24 (2.5%) | 374 (23.8%) |
| Hypertension | 550 (57.6%) | 644 (41.0%) |
| Diabetes mellitus | 304 (31.8%) | 366 (23.3%) |
| 25 ≤ BMI < 30 | 324 (33.9%) | 477 (30.3%) |
| BMI≥30 | 223 (23.4%) | 392 (24.9%) |
| Coronary Artery Disease | 343 (35.9%) | 313 (19.0%) |
| Left Ventricular Ejection Fraction (LVEF) | 59.7% ± 11.6% | 60.8% ± 9.5% |

**Figure 2.**
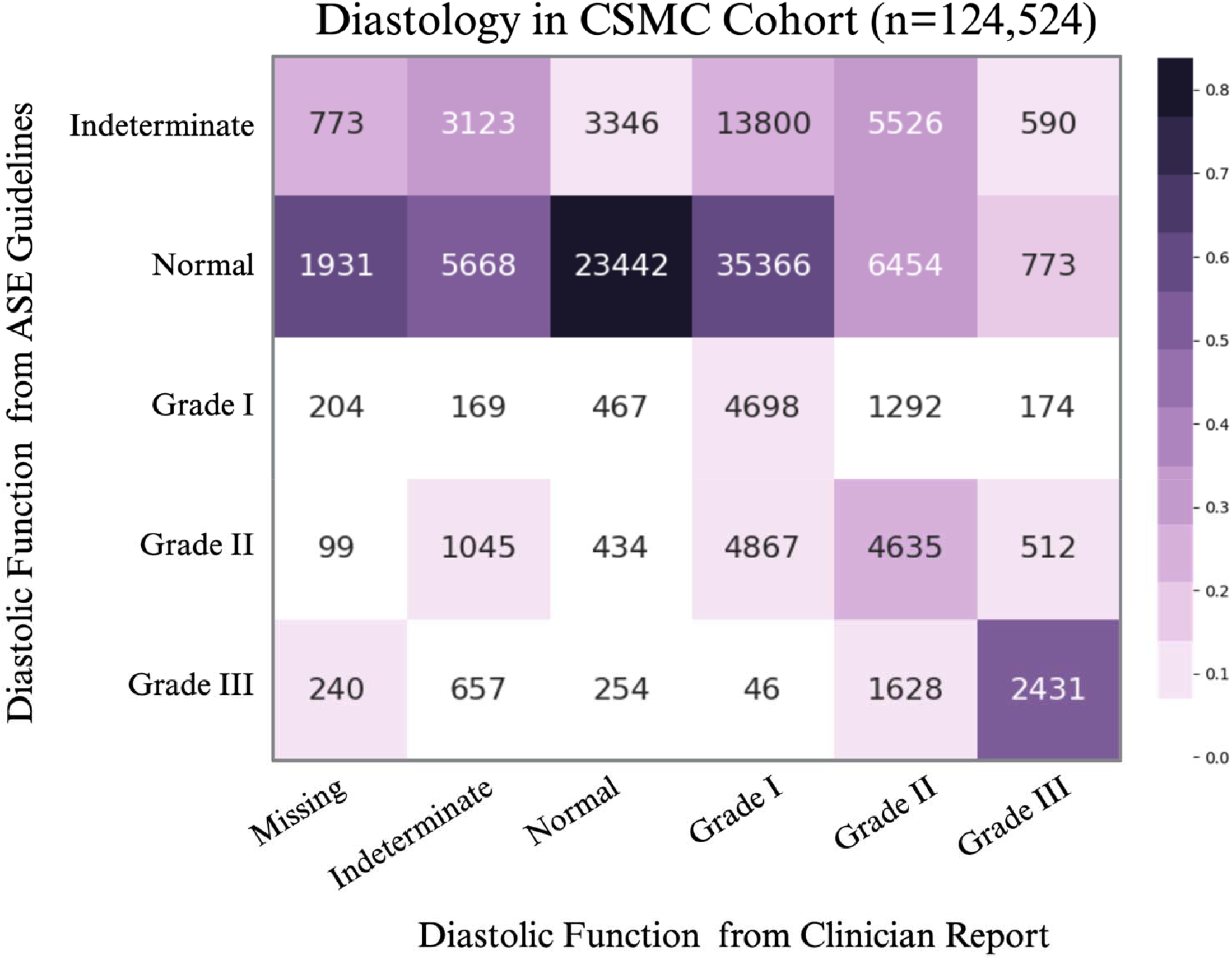
Confusion matrix of diastolic function extracted from clinician reports versus diastolic function calculated from clinical measurements for 87,425 patients who received care at CSMC from April 2016 to November 2024

**Figure 3.**
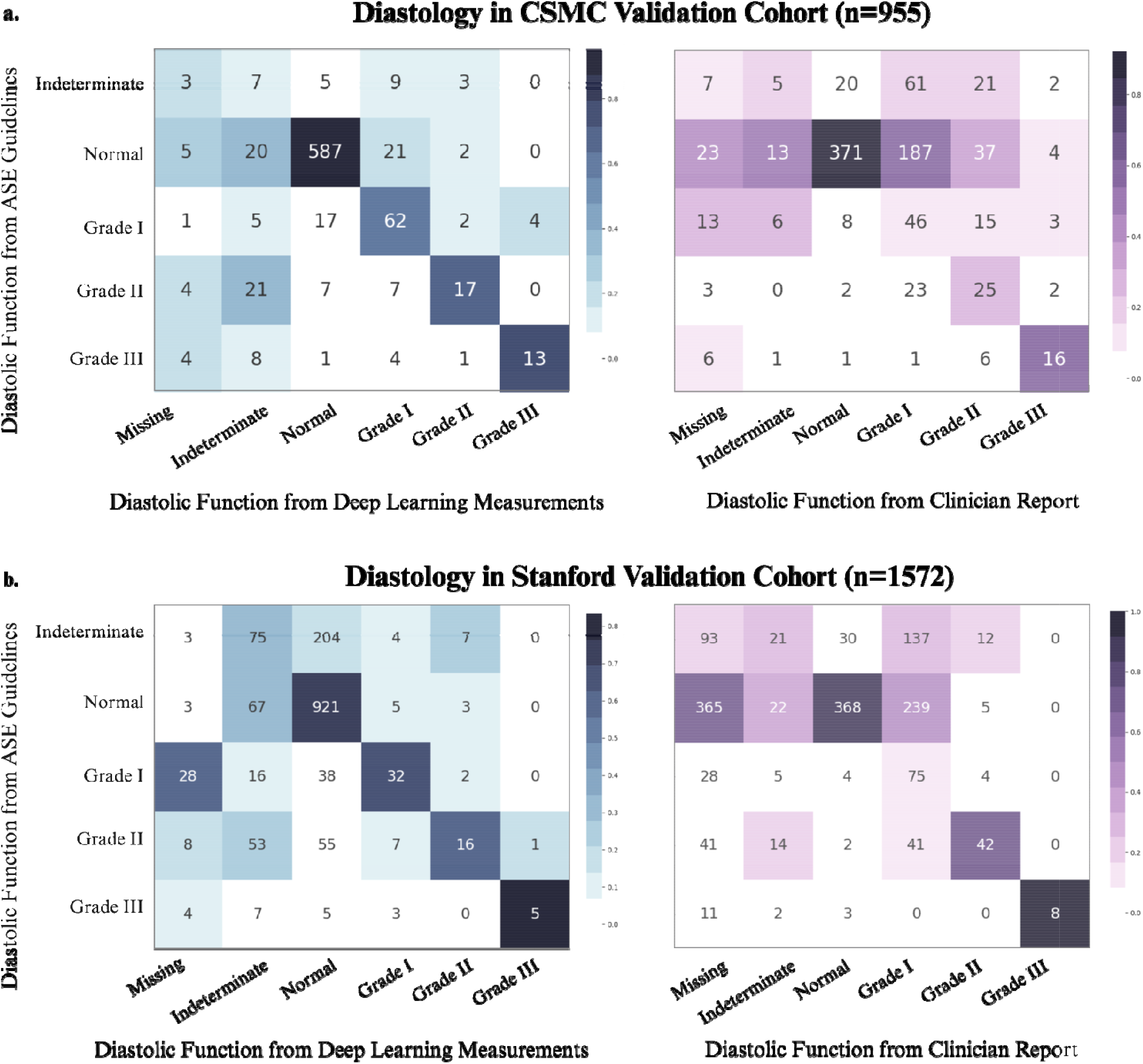
Confusion matrix of diastolic function derived from ASE guidelines versus DL measurements (left) and ASE guidelines versus clinician report (right) in the CSMC validation cohort (top) and the SHC validation cohort (bottom)

In the CSMC cohort, 16.3% of patients were missing a clinician assessment of diastolic function; of these 20,355 studies, 16.0% of these studies had some degree of diastolic dysfunction when using ASE 2016 criteria. In the SHC cohort, 34.2% of studies are missing a clinician assessment, with 14.9% having some grade of diastolic dysfunction. Additionally, clinicians reported a wide range of diastology for patients who had indeterminate function by ASE guidelines (Figure 2). There was not much heterogeneity by physician or patient subgroups (Table 2, Supplemental Figure 1).

**Table 2.** Agreement across patient subgroups for clinician report and DL diastology with ASE guidelines in the CSMC and SHC cohorts.

| Subgroup | Agreement between ASE Guidelines and Clinician Report |  | Agreement between ASE Guidelines and DL Measurement |  |
| --- | --- | --- | --- | --- |
|  | CSMC | SHC | CSMC | SHC |
| Female | 18.2% | 34.5% | 79.7% | 68.7% |
| Male | 16.8% | 30.9% | 73.8% | 64.7% |
| Age <65 | 25.3% | 42.2% | 85.6% | 78.3% |
| Age ≥65 | 12.8% | 20.2% | 69.3% | 51.6% |
| Hypertension | 14.1% | 22.7% | 69.4% | 56.4% |
| Diabetes mellitus | 15.1% | 28.7% | 80.8% | 61.2% |
| BMI ≥ 30 | 28.1% | 32.6% | 79.7% | 53.0% |
| Coronary Artery Disease | 14.5% | 31.6% | 69.2% | 66.3% |

### Deep Learning Pipeline to Automate Diastology

Our automated deep learning pipeline performs comprehensive assessment of full transthoracic studies, including view classification^26^, assessment of left ventricular ejection fraction^16^, and automated measurements of Doppler parameters^19^. In two held out patient cohorts not used for model development, we evaluated the performance of our deep learning pipeline. The CSMC validation cohort consisted of 955 studies across 943 unique patients who received care at CSMC from June 2022 to November 2024 (Table 1). The SHC validation cohort consisted of 1,572 studies across 1,560 patients. Our model demonstrated strong agreement of 76.5% and 66.7% in CSMC and SHC, respectively (Figure 3, Supplemental Figures 2, 3, and 4). Weighted Cohen’s kappa between DL measurements and ASE guidelines was 0.52 and 0.27 in the CSMC and SHC cohorts, respectively, versus kappa of 0.29 and 0.06 between clinician evaluation and ASE guidelines in CSMC and SHC, respectively.

Agreement between DL measurements and ASE guidelines was consistently strong across patient subgroups based on sex, age, hypertension, diabetes mellitus, obesity, and coronary artery disease (Supplemental Figure 5). In CSMC, DL measurements had agreement ranging from 69-85% and in SHC, from 51-78.5% (Table 2). There were 255 discordant cases between DL measurements and ASE guidelines, with weak correlations for abnormal LAV_i_, E/e’ ratio, and E/A ratio, and E velocity; of the 523 discordant cases in the SHC cohort, DL predicted values for medial e’ velocity and abnormal LAV_i_ were the weakest (Supplemental Table 2). DL diastology correctly identified patients with elevated LV filling pressure with an area under the receiver operating curve (AUROC) of 0.86 and 0.73 in CSMC and SHC, respectively.

## Discussion

In this work, we evaluated the precision of clinical assessment of diastolic function and introduce a deep learning-based pipeline that automates the clinical workflow to characterize diastolic function from echocardiography. Our models demonstrated strong performance in two distinct health care centers and across a variety of patient subgroups. In combination, the DL measurements have nearly 30% stronger agreement with ASE guidelines than holistic clinician evaluation.

LVDD underlies a range of cardiac pathologies, such as HF, valvular disease, and cardiomyopathy, in addition to extracardiac pathologies such as renal disease and diabetes mellitus. In this work, we highlight the discordance between clinician evaluation of diastology with clinical guidelines in two large academic centers. Our analysis reinforces the need for efficient, reproducible assessment of diastology as imprecision in diastology can contribute to missed diagnoses. To this end, we developed a DL pipeline that automates the grading of diastolic function. Our workflow directly addresses major bottlenecks to routine clinical diastology – time-intensity and variability. DL-diastology demonstrated increased concordance with ASE guidelines and can facilitate precision phenotyping of diastolic function with higher accuracy and reproducibility than current clinical practice.

Limitations of our study include the study of echocardiographic images from academic tertiary care centers, which may bias patient selection. Model performance should be further examined in care centers across the country. Future work includes correlation of DL diastology with cardiovascular outcomes and comparing sonographer to model performance in real-time.

## Conclusion

We developed a deep learning pipeline to automate the clinical workflow for assessing diastology. Our findings demonstrate that automated deep learning diastology has higher agreement with ASE guidelines than holistic clinician evaluations. Our deep learning workflow can increase the efficiency, completeness, and precision of diastology and contribute to precision phenotyping of diastolic dysfunction.

## Supporting information

Supp Results

## Data Availability

All data produced in the present work are are available upon reasonable request to the authors

