## Supplementary material for "Automated Deep Learning Pipeline for Characterizing Left Ventricular Diastolic Function": Supp Results

**Supplemental Information**

**Supplemental Figure 1.** Agreement of diastology between clinician report and echocardiographic measurements by ASE guidelines for physicians at CSMC; 123,459 studies were read by 32 physicians.

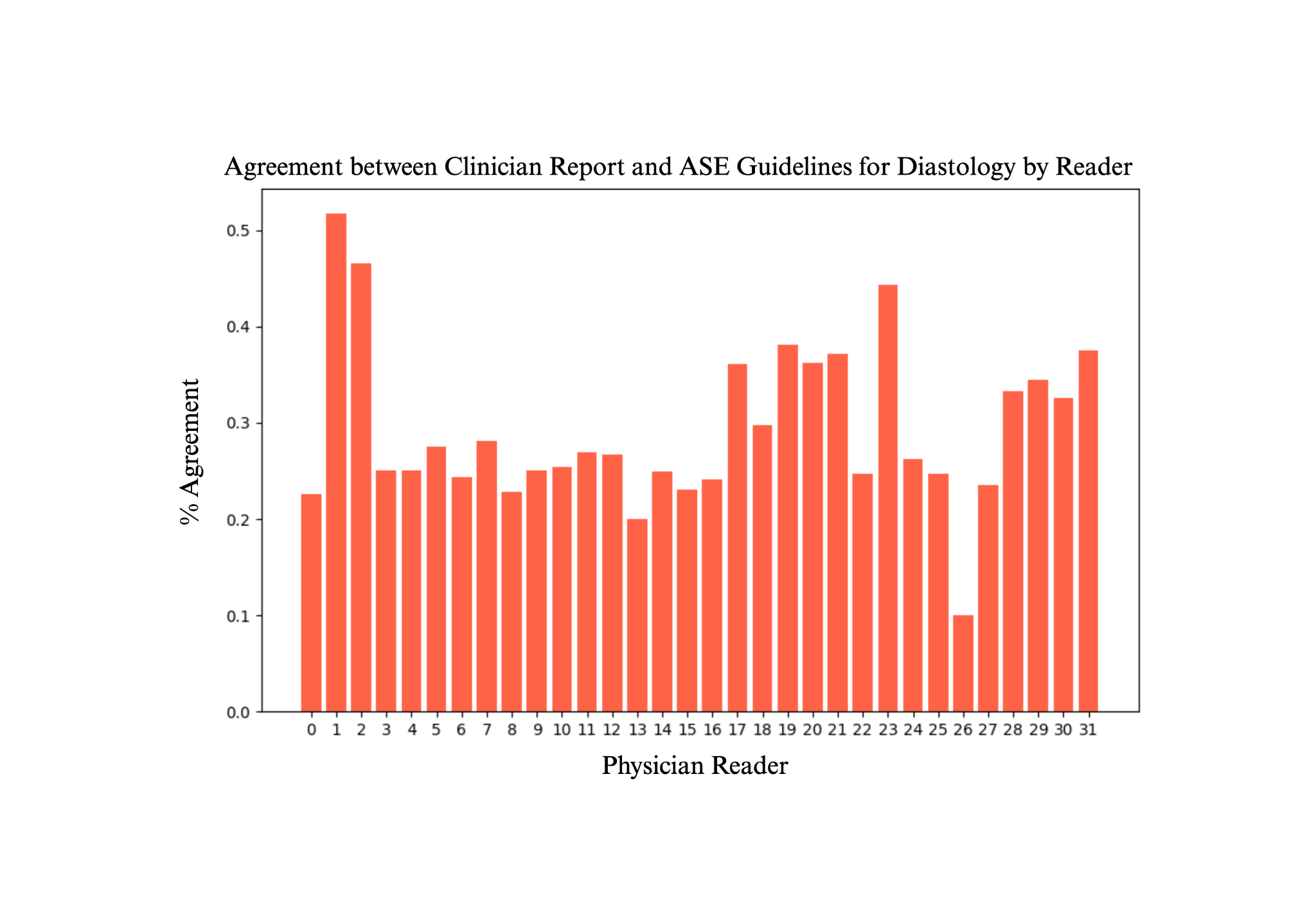

**Supplemental Figure 2.** Correlation between clinical measurements and DL measurements to characterize diastolic function in the CSMC 2022-2024 cohort

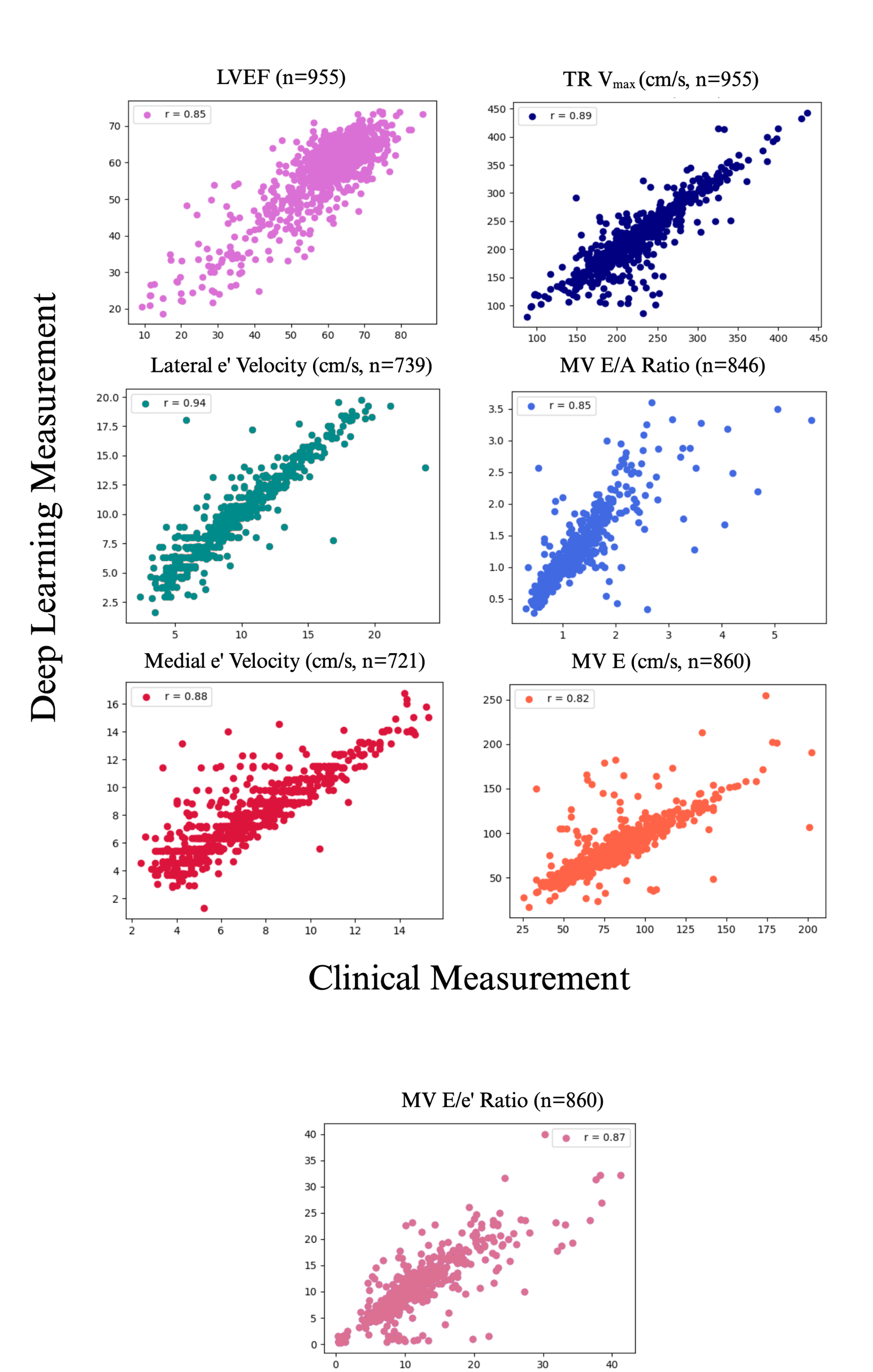

**Supplemental Figure 3.** Correlation between clinical measurements and DL measurements to characterize diastolic function in the SHC cohort

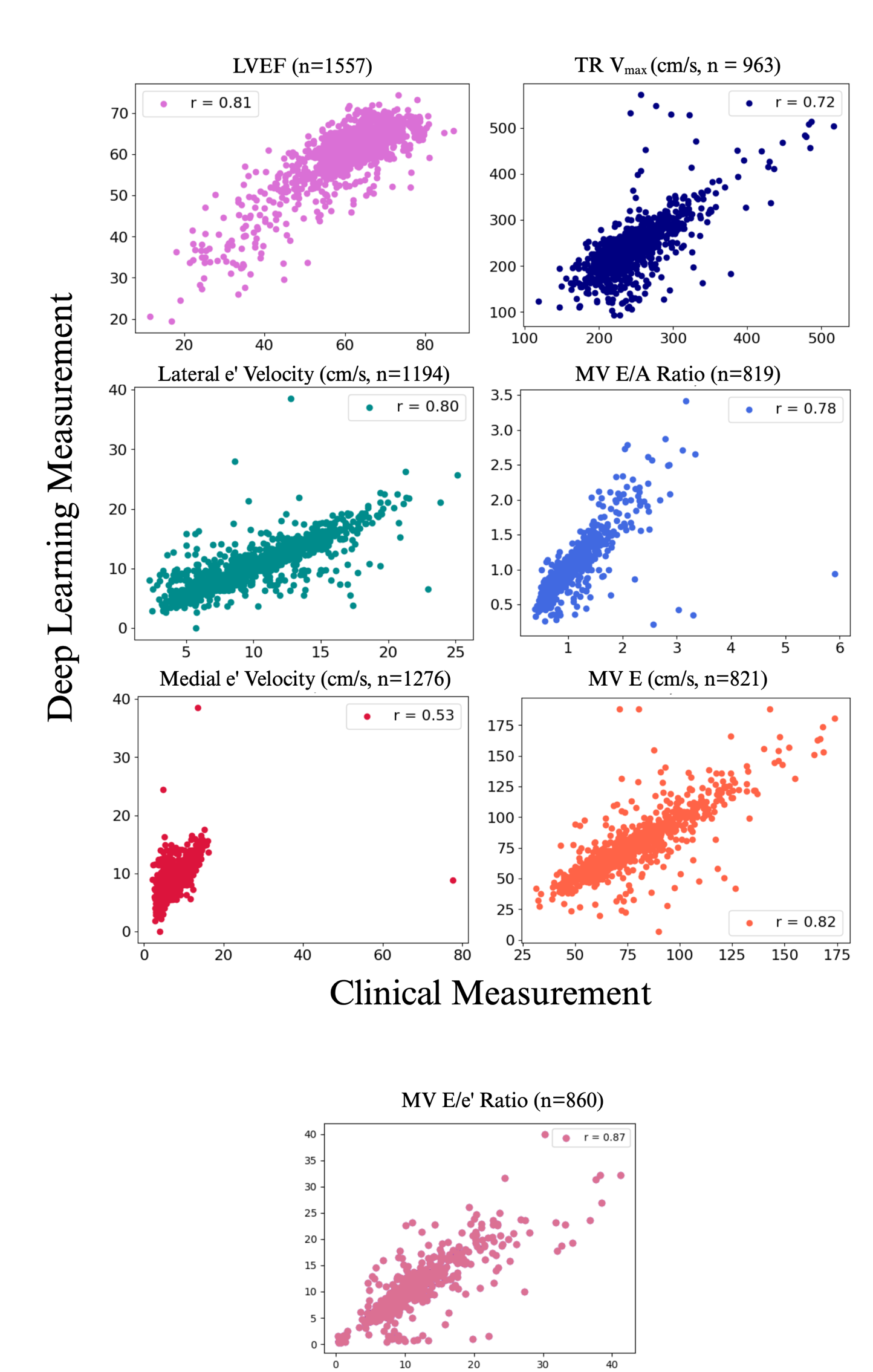

**Supplemental Figure 4.** Representative studies with concordance between clinical and DL measurements

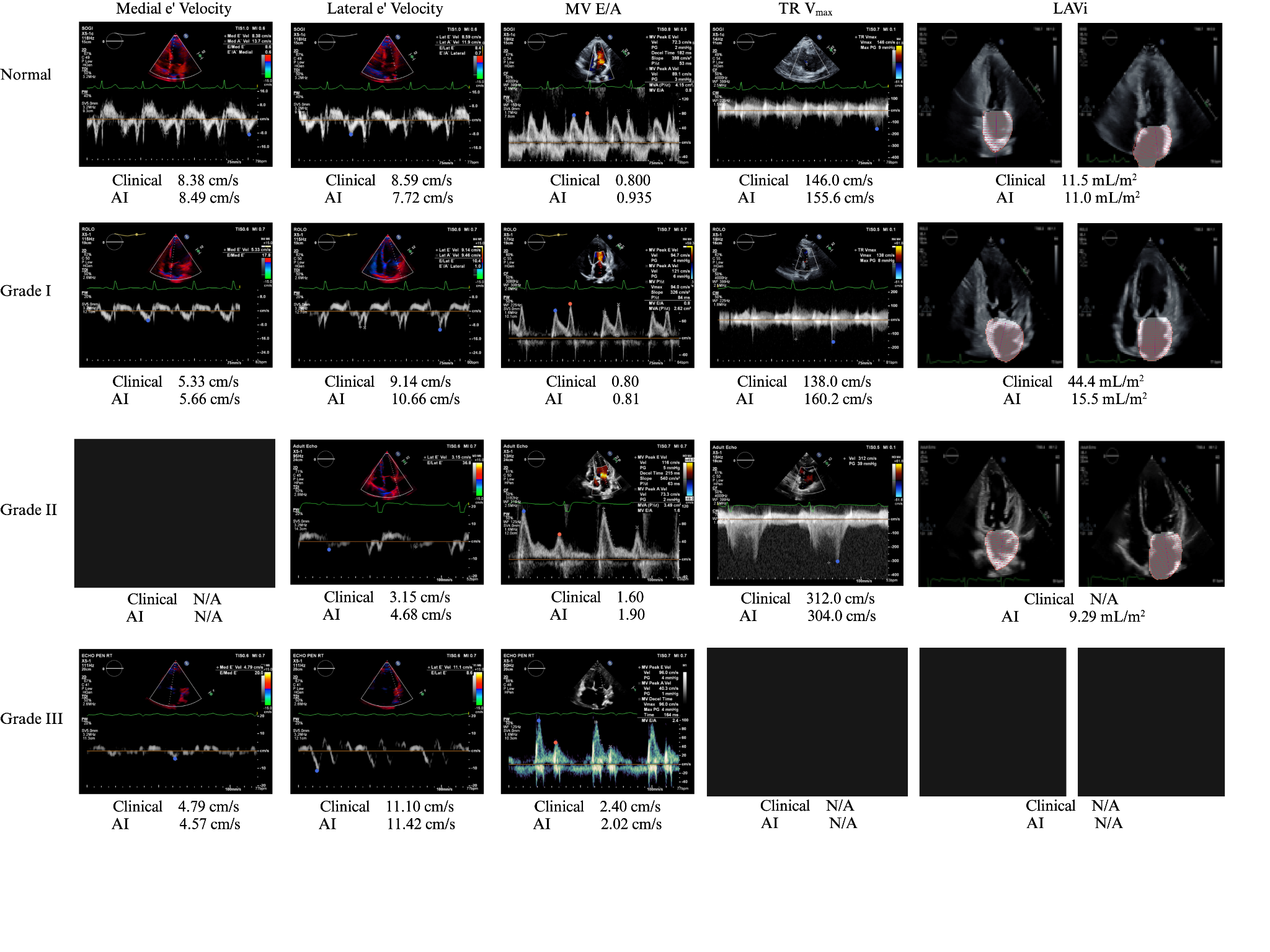

**Supplemental Figure 5.** Diastolic function from clinician reports, clinical measurements, and DL measurements in patients with hypertension (htn), diabetes mellitus (DM), obesity, and coronary artery disease (CAD) in the CSMC validation cohort (a-d) and the SHC validation cohort (e-h).

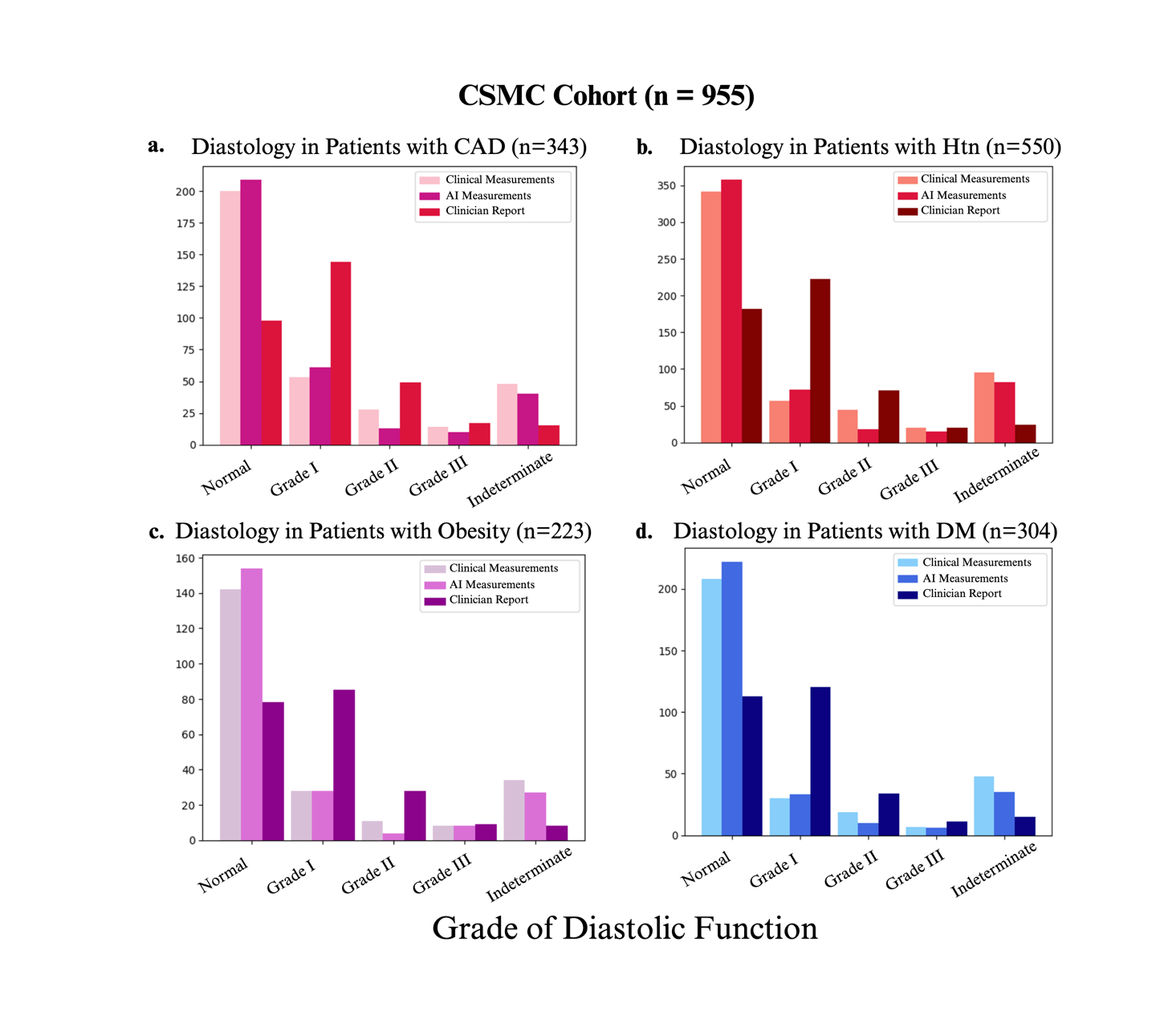

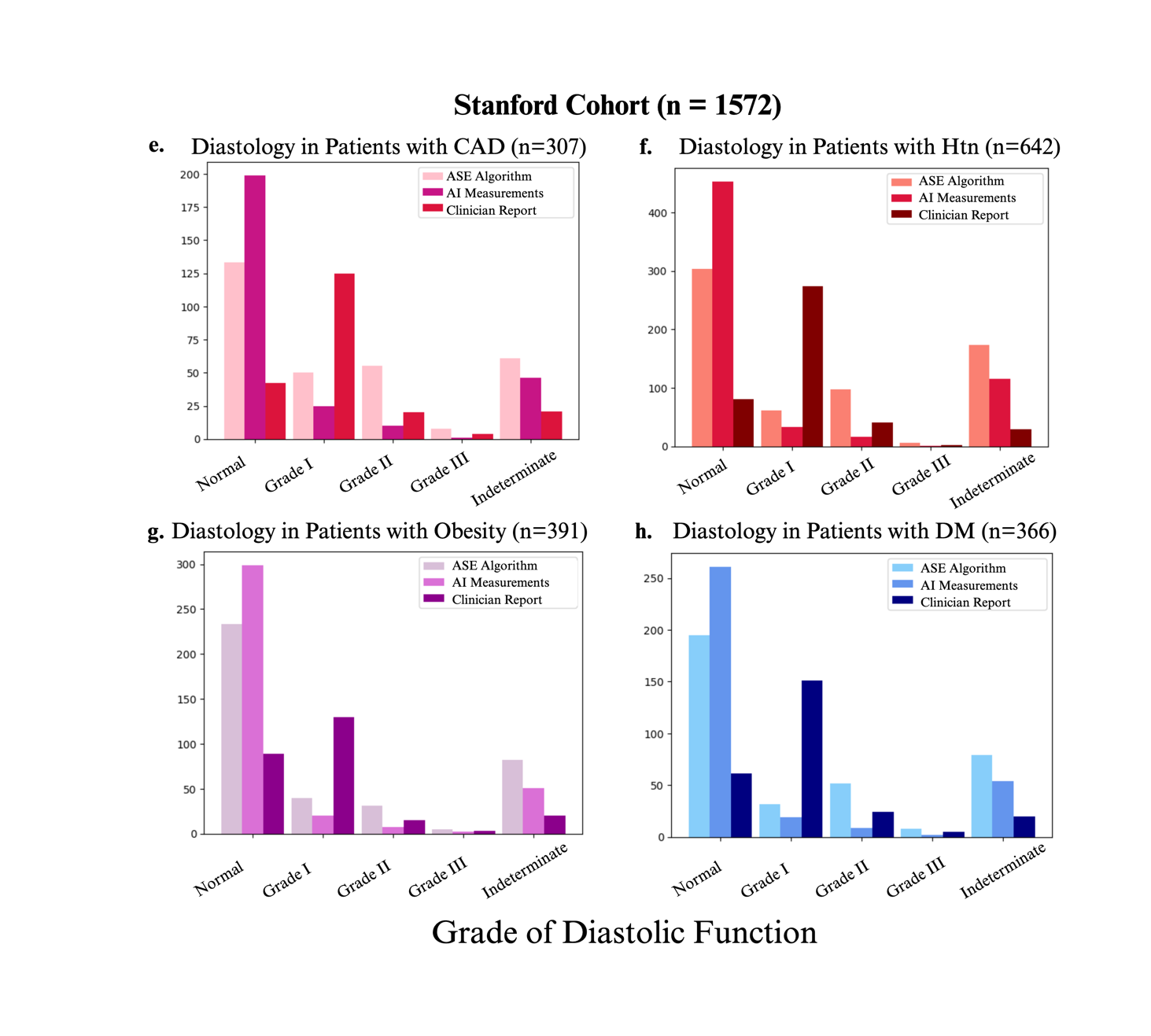

**Supplemental Table 1.** Characteristics of patients in CSMC Historical Echo cohort from April 2016 to June 2022

| **Characteristic** | **n (%)** |
| --- | --- |
| Female | 33,281 (47.4%) |
| Male | 36,961 (52.6%) |
| Age (mean±std) | 64.7 ± 17.3 |
| Race and Ethnicity | |
| American Indian / Alaskan Native | 173 (0.23%) |
| Asian | 5,130 (7.3%) |
| Black / African-American | 9,884 (14.1%) |
| Hispanic | 9,033 (12.9%) |
| Native Hawaiian / Pacific Islander | 220 (0.31%) |
| White | 48,161 (68.6%) |
| Unknown | 1,521 (2.2%) |
| Hypertension | 32,670 (46.5%) |
| Diabetes mellitus | 20,431 (20.8%) |
| BMI < 25 | 1113 (1.6%) |
| 25 ≤ BMI < 30 | 149 (0.2%) |
| BMI≥30 | 61,378 (87.2%) |
| Coronary Artery Disease | 18,562 (26.4%) |
| Left Ventricular Ejection Fraction (LVEF) | 60.2% ± 12.0% |

**Supplemental Table 2.** Correlation between clinical measurements and DL measurements in discordant cases in the CSMC and SHC validation cohorts

| **Parameter** | **Pearson r Coefficient** | |
| --- | --- | --- |
|  | **CSMC** | **SHC** |
| LVEF | 0.81 | 0.85 |
| TR V_max_ | 0.98 | 0.70 |
| Medial e’ velocity | 0.91 | 0.38 |
| Lateral e’ velocity | 0.98 | 0.65 |
| E velocity | 0.34 | 0.84 |
| E/e’ ratio | 0.51 | 0.60 |
| E/A ratio | 0.25 | 0.67 |
| Abnormal LAV_i_ | -0.10 | -0.17 |
